# Integrating immunophenotypes reveals causal relationship between tuberculosis and lung cancer: Mendelian randomization study

**DOI:** 10.1101/2024.06.10.24308725

**Authors:** Xianwen Li, Dayin Huang, Chang Liu, Wei Yu, Lijun Bi

## Abstract

**Background:** Tuberculosis (TB) and lung cancer (LC) are major respiratory diseases causing significant deaths worldwide. Both primarily affect the lungs and share similar clinical symptoms. The comorbidity rate between TB and LC is high, and epidemiological data suggest a potential association between the two diseases. However, the detailed causal relationships and potential mediators remain underexplored due to limited evidence from observational studies. This study aims to elucidate the relationship between TB and LC, including the pathological subtypes of LC, and to investigate the potential mediating effects of immune cells.

**Materials and Methods:** The genetic variants for TB and LC were sourced from the IEU Open GWAS Project, while the data for the 731 immunocyte phenotypes were obtained from their genome-wide association studies (GWAS). The inverse variance-weighted (IVW), MR-Egger, weighted median, simple mode and weighted mode were used to evaluate the causal relationship between TB and LC. We then applied a two-step Mendelian randomization (MR) analysis to examine the roles of immunocyte phenotypes as mediators. Additionally, multiple sensitivity analyses were conducted to ensure the reliability of the MR results.

**Results:** The results of the IVW methods of the MR analysis indicated a causal relationship between TB and LC, demonstrating a positive correlation (OR = 1.072, 95% CI: 1.010–1.137, *P* < 0.05). The immune phenotype CD4 on CD39+ resting regulatory T cell is implicated in the linkage between TB and LC as well (mediation proportion: 9.09%). Additionally, no evidence was found to support a reverse causal relationship between TB and LC, nor a causal relationship between TB and the pathological subtypes of LC. There was no observed heterogeneity or horizontal pleiotropy in our MR results.

**Conclusion:** Our study suggests the causal effect of TB on increased LC risk, with immune phenotype playing a mediating role. These insights provide a significant basis for the prevention and concurrent management of LC and TB.

## Introduction

Tuberculosis (TB) and lung cancer (LC) are significant respiratory diseases that pose a major threat to human health. TB, caused by *Mycobacterium tuberculosis* (MTB), is a major global health challenge, with 10.6 million cases and 1.3 million deaths in 2022[1]. According to the latest International Agency for Research on Cancer (IARC) data, LC remains the most prevalent malignancy globally, with 2.48 million new cases in 2022 and causing 1.8 million deaths annually[2]. LC are divided into non-small cell lung carcinoma (NSCLC) and small cell lung carcinoma (SCLC). NSCLC includes adenocarcinoma (LUAD), squamous cell carcinoma (LUSC), and large cell carcinoma (LCC)[3]. The co-existence of TB and LC has been well-documented in the literature, spanning from longitudinal studies to cross-sectional epidemiological reports.

Epidemiologic data suggest that a statistically significant association between TB and LC. For example, a meta-analysis of 16 studies demonstrated that patients with a prior TB diagnosis have an elevated risk of developing LC (RR = 1.48, 95% CI: 1.17, 1.87; *I^2^*= 54.27%)[4]. Another comprehensive nationwide cohort study reported a 1.67-fold increased risk of LC in TB patients compared to non-TB individuals[5]. Moreover, it has been reported that the incidence rate of TB among LC patients is increasing, with an incomplete treatment rate for TB in these patients reaching 61.13%[6]. Even after the completion of TB treatment, the recurrence rate of TB in malignant tumor patients within two years is as high as 5.03%[7]. Moreover, reports indicated a higher incidence of irritable cough and hemorrhagic pleural effusion in TB patients with LC compared to those with TB alone[8]. However, associations derived from observational studies may be affected by reverse causality and confounding factors. Therefore, whether TB is causally associated with LC risk remains unknown[9]. Further research is required to support the causal association between TB and LC.

While immunity plays a crucial role in the pathogenesis of TB and LC, emerging evidence underscores the importance of genetic characterization in understanding these diseases, thereby offering a foundation for their prevention and treatment[10,11]. Mendelian randomization (MR) leverages genome-wide association studies (GWAS) to use genetic variants as instrumental variables (IVs), enabling the inference of causal effects while minimizing confounding factors[12]. In the present study, we employed MR as a methodological framework to ascertain the potential causal nexus between TB and LC. Then, a two-step MR approach was used to investigate the mediation effect of 731 immunophenotypes in the association between TB and LC.

## Materials and methods

### Study design

This bidirectional two-sample MR study utilized publicly available GWAS datasets and adhered to three essential MR assumptions[13]: strong association of IVs with the exposure, independence of IVs from confounders, and exclusive influence of IVs on the outcome through the exposure. TB was designated as the exposure and LC as the outcome, employing Single Nucleotide Polymorphism (SNP) significantly associated with TB as IVs. We also performed a mediation analysis to explore potential pathways between TB and LC (Figure 1). Ethical approval was not required as data were sourced from public databases.

**Figure 1.**
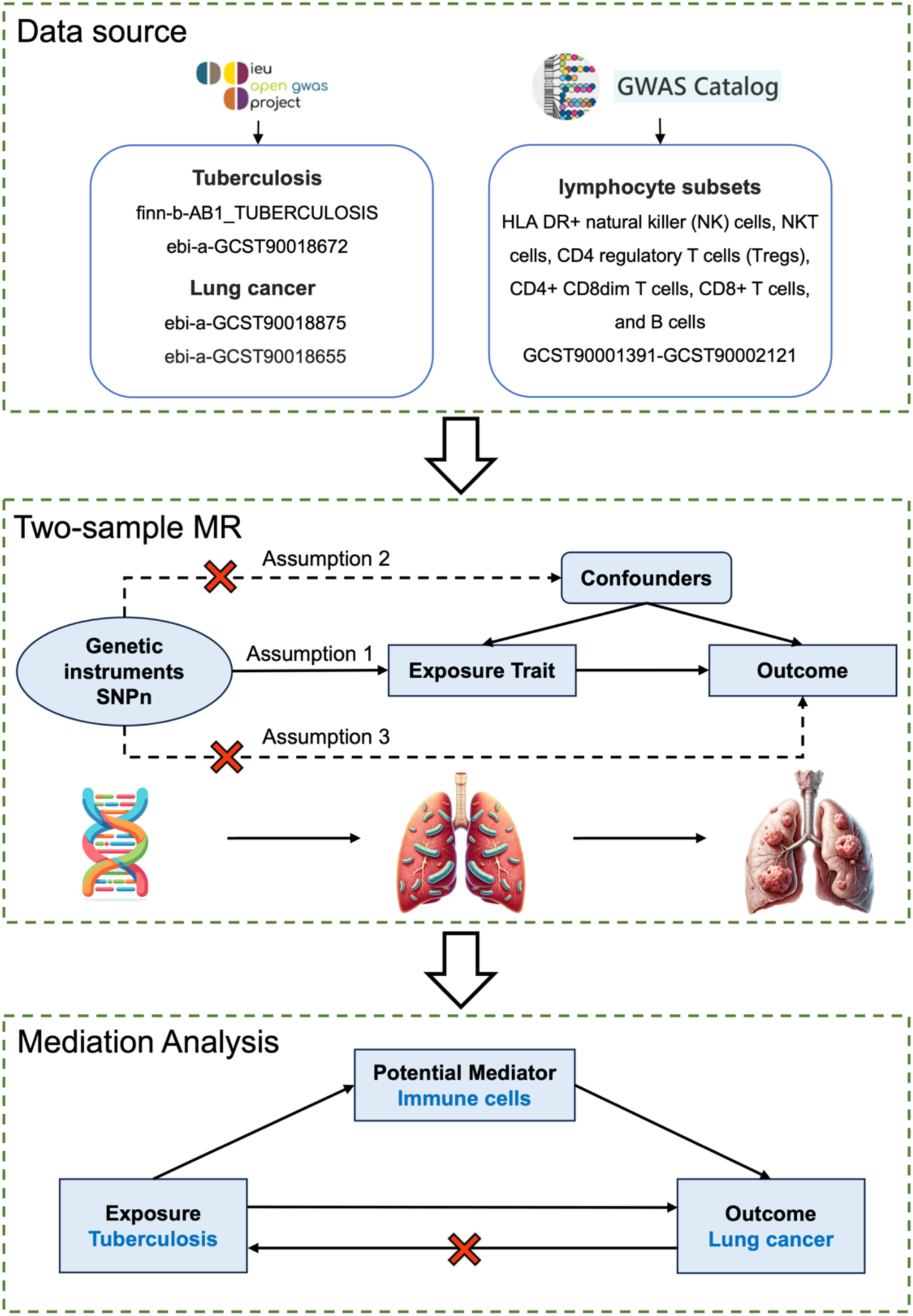
Overview of MR assumptions. MR study rationale: assumption 1, Genetic instruments must be associated with the exposure.; assumption 2, Genetic instruments must not be associated with any confounders.; assumption 3, Genetic instruments must not be directly associated with the outcome; they should influence the outcome solely through their impact on the exposure.

### Data sources

The genetic variants for LC and TB were obtained from the IEU Open GWAS Project (https://gwas.mrcieu.ac.uk/). The GWAS ID for TB are finn-b-AB1_TUBERCULOSIS and ebi-a-GCST90018672, while the ID for LC are ebi-a-GCST90018875 and ebi-a-GCST90018655. For distinct pathological subtypes of LC, the LUAD relevant GWAS identifier are ebi-a-GCST004744 and ieu-a-984, the LUSC relevant GWAS identifier are ebi-a-GCST004750, ieu-a-967 and ieu-a-989 and for SCLC, the ID is ebi-a-GCST004746. The significance level for each IV was set at 5 × 10^8^. Since no SNP were selected as genetic instruments for TB that reached genome-wide significance, we adopted a less stringent threshold *P* < 5 × 10^6^ to identify more SNPs for TB in finn-b-AB1_TUBERCULOSIS. We excluded correlated SNPs at a linkage disequilibrium threshold of r^2^ > 0.01, retaining SNPs with the strongest effect. The population in the above datasets included the Europeans and Asians.

Comprehensive information on each immunophenotype in peripheral blood can be publicly accessed from the GWAS Catalog (accession numbers from GCST90001391 to GCST90002121, including 4 trait types, such as 118 absolute counts (AC), 389 median fluorescence intensity (MFI), 32 morphological parameters (MP) and 192 relative counts (RC). These features encompass various developmental stages and cell types of immune cells. The significance level for IVs for each immunophenotype was set at 1×10^5^[14]. We pruned these SNPs (linkage disequilibrium [LD] r^2^ threshold of <0.001 within a 10000 kb distance).

### Instrumental variable selection

In this study, IVs were selected based on stringent criteria: 1) IVs were chosen at the locus-wide significance level of *P* < 1 × 10^5^ if insufficient genome-wide significant loci were available, or at a genome-wide significance threshold of *P* < 5 × 10^8^ for each exposure trait. 2) SNPs associated with the outcome at *P* < 0.05 were excluded. 3) Linkage disequilibrium was managed through clumping (r^2^ < 0.01, window size = 5000 kb; or r^2^ < 0.001, window size = 10,000 kb) to minimize bias. 4) Horizontal pleiotropy was assessed using the MR-PRESSO test, removing outliers to address pleiotropy (P-value for MR-PRESSO global test > 0.05). 5) The strength of IVs was determined using an F-statistic; those with an F-statistic < 10 were excluded to mitigate weak instrument bias, calculated as F = [R^2^ × (n − k − 1)] / [k × (1 − R^2^)], where R^2^ represents the variance in exposure explained by IVs, n is the sample size, and k the number of IVs. 6) IVs directly related to outcomes were eliminated.

### Statistical analysis

#### Two-sample Mendelian randomization

To elucidate the causal dynamics between TB and LC, we applied MR methods, assessing the influence of TB on LC development. For single IV exposure, the Wald ratio was utilized to infer causality. For exposures comprising multiple IVs, a comprehensive array of methods was employed, including inverse variance-weighted (IVW), simple mode, MR-Egger, weighted median, and weighted mode. IVW, favored for its high statistical power[15], was primarily used, as it integrates the Wald ratio estimates of each IV into a meta-analysis framework with the intercept set to zero. This method is particularly effective in the absence of horizontal pleiotropy, providing unbiased causal estimates[16].

When heterogeneity was present, a random-effect IVW test was applied to provide more conservative and robust estimates; a fixed-effect model was used otherwise. Although the simple mode method is not as powerful as the IVW method, it provides robustness against pleiotropy[17]. The MR-Egger method was employed to detect and account for multiple horizontal effects, providing valid causal estimation even in the presence of pleiotropy[18]. The weighted median could deliver reliable causal estimates even when up to 50% of the IVs were ineffective[19], and the weighted mode approach remained valid provided the majority of IVs with similar causal estimates were sound[20].

To ensure the robustness of our findings, sensitivity analyses including MR-Egger regression and MR-PRESSO were conducted to assess horizontal pleiotropy. A non-zero intercept in the MR-Egger regression indicated directional pleiotropy[18]. Cochran’s Q test assessed the heterogeneity among IVs, while leave-one-out (LOO) sensitivity analysis explored the impact of individual SNPs on the causal estimation. Only findings showing no significant heterogeneity or pleiotropy and achieving a significance threshold of *P* < 0.05 in the IVW MR method were considered robust and included in the final analysis.

#### Reverse Mendelian randomization analysis

To assess whether LC has a causal effect on TB (IVW, *P* < 0.05), we conducted a reverse MR analysis. In this analysis, SNPs associated with LC served were utilized as IVs, with LC positioned as the exposure and TB as the outcomes. The procedure for the reverse MR mirrored that of the standard MR analysis.

#### Mediation analysis

For significant associations, potential mediation effects (the exposure– mediator–outcome pathway) may exist. The mediation analysis in this study focused on immunophenotype. Initially, multivariable MR (MVMR) was used to screen immunophenotype that had a causal relationship with LC to obtain beta (B). Subsequently, the causal association between TB and immunophenotype was assessed using two-sample MR methods after correction for LC to obtain beta (A) and ensure that the mediating effects on outcomes are independent of exposure[21]. The mediation effect was quantified using a two-step MR approach: mediation effect = beta (A) × beta (B). The total effect of the TB on LC was obtained in the previous two-sample MR, and direct effect = (total effect − mediation effect). The mediation proportion used the following formula: mediation proportion = (mediation effect / total effect) × 100%. The 95% confidence intervals (CI) for the mediation effects and proportions mediated were estimated using the delta method[21]. When a triangular relationship existed, representing that exposure was causally associated with outcome, mediator was causally associated with outcome, and exposure was causally associated with mediator. The identified immunophenotype were considered to have potential mediation effects in the pathway from TB to LC.

MR analyses were performed using the R packages ‘TwoSampleMR’, ‘MendelianRandomization’ and ‘RMediation’. All results are reported as ORs with 95% CIs. All statistical analyses were performed in R software (v.4.3.2).

## Results

### Exploring the Causal Relationship Between TB and LC

In this study, we evaluated the potential causal association between TB and LC using two-sample MR. Our analysis primarily used the IVW method, complemented by MR-Egger, weighted median, simple mode and weighted mode. The IVW analysis suggested that TB likely increases the risk of LC, as indicated by an OR of 1.072 with a 95% CI of 1.010–1.137, and a statistically significant P-value of less than 0.05 (Figure 2, Table S1).

**Figure 2.**
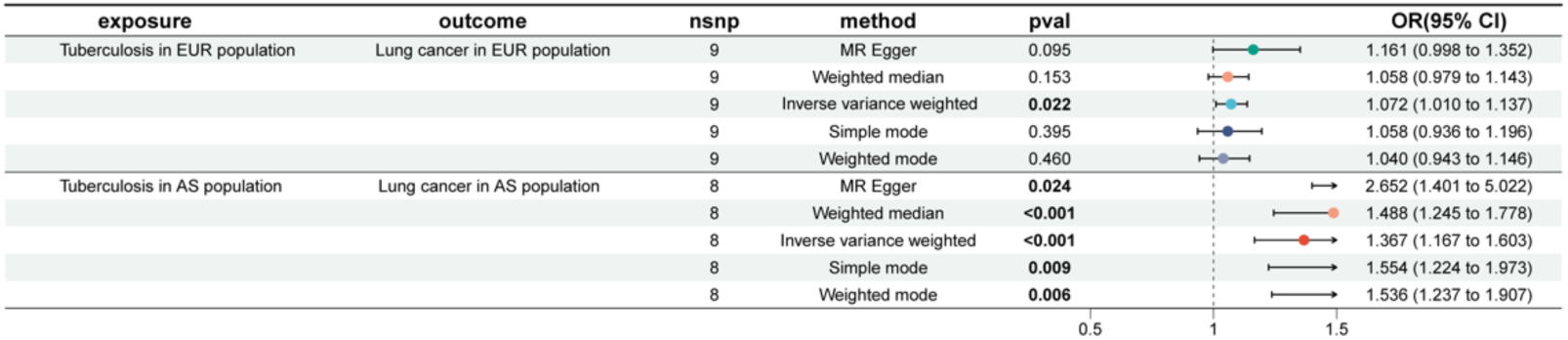
MR analyses of TB and LC. MR analyses demonstrate the causal effect of TB on LC in European and Asian populations using five methods: MR Egger, weighted median, IVW, simple mode, and weighted mode. OR, 95% CI, and *P*-values are shown for each method.

To ensure the robustness and reliability of our findings, comprehensive validations were performed, including tests for horizontal pleiotropy, heterogeneity assessments, and LOO analyses. The evaluation of horizontal pleiotropy via the MR-Egger method revealed a regression intercept P-value of 0.297, indicating no significant pleiotropic effects (Table S2). Additionally, the LOO analysis underscored consistent causal estimates across all included SNPs (Figure S1). Scatter plots further substantiate the robustness and consistency of the causal associations observed (Figure S2).

Asia remains a high-incidence region for TB, with both the absolute number of cases and the severity of the disease markedly elevated compared to global averages[1]. We further assessed the potential causal relationship between TB and LC within East Asian populations. Employing the IVW method, alongside MR-Egger, weighted mode, simple mode and weighted median approaches, all five methods consistently identified TB as a risk factor for LC (Figure 2 and S3). Specifically, the IVW analysis revealed that the incidence of LC in individuals with TB was 1.367-fold higher than those without TB (OR = 1.367, 95% CI: 1.167-1.603, *P* < 0.001). And the plot of LOO analysis is indicated reliable data (Figure S4). The MR findings were further corroborated by sensitivity analyses, which consistently indicated the absence of heterogeneity and horizontal pleiotropy (Table S3). Subsequently, we employed MR to further analyze the causal relationship between TB and various pathological subtypes of LC. The results showed no statistically significant differences (Figure 3).

**Figure 3.**
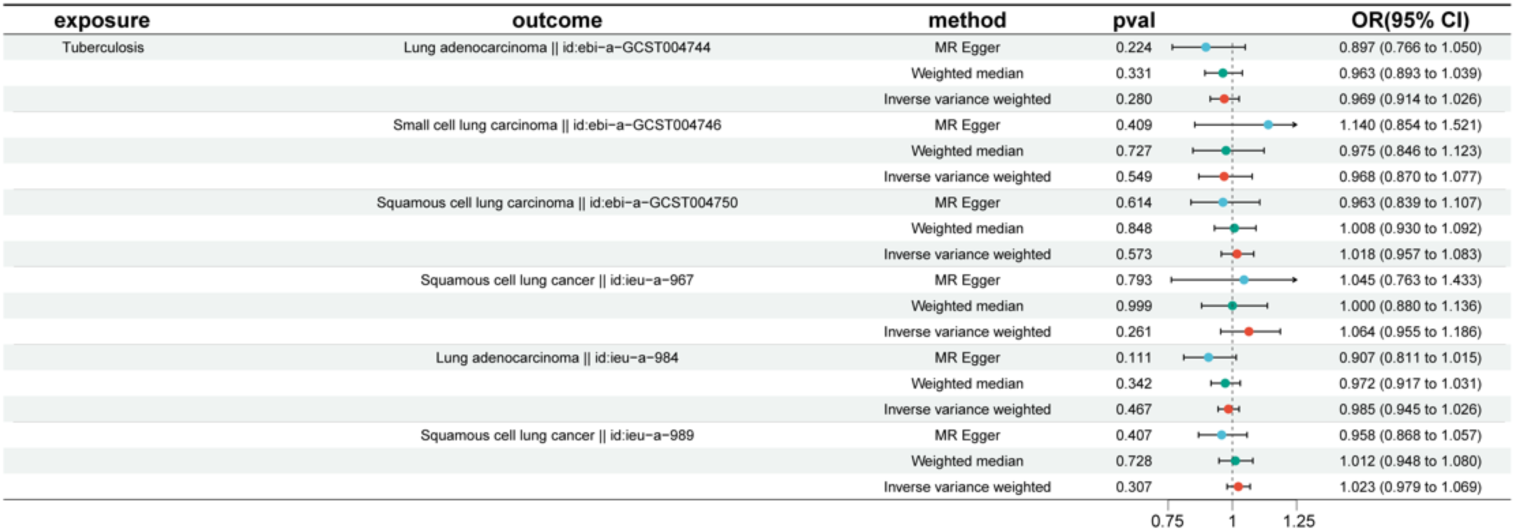
MR analyses of TB and LC with different pathological subtypes. Analyses utilized three methods: MR Egger, weighted median, and IVW. OR with 95% CI and *P*-values are presented for each method. The results showed no statistically significant difference.

### Bidirectional Causal Relationships Between TB and LC

To evaluate reverse causality between TB and LC, we treated overall LC as the exposures with TB as the outcome. Ultimately, our findings show no evidence of a reverse causal relationship between TB and LC (Table S4-5).

### Mediation analysis results

To explore the potential immunological mechanisms underlying the association between TB and LC, we executed a two-step mediation analysis to trace the causal pathways from TB to LC. This investigation was extended through a MVMR study, which utilized 731 immune cell phenotypes as exposures, identifying 30 phenotypes that are significantly associated with LC (*P* < 0.05) (Figure 4 and Table S6). The MR results demonstrated stability across the sensitivity analyses, indicating no significant heterogeneity or horizontal pleiotropy (Table S7 and Figure S5).

**Figure 4.**
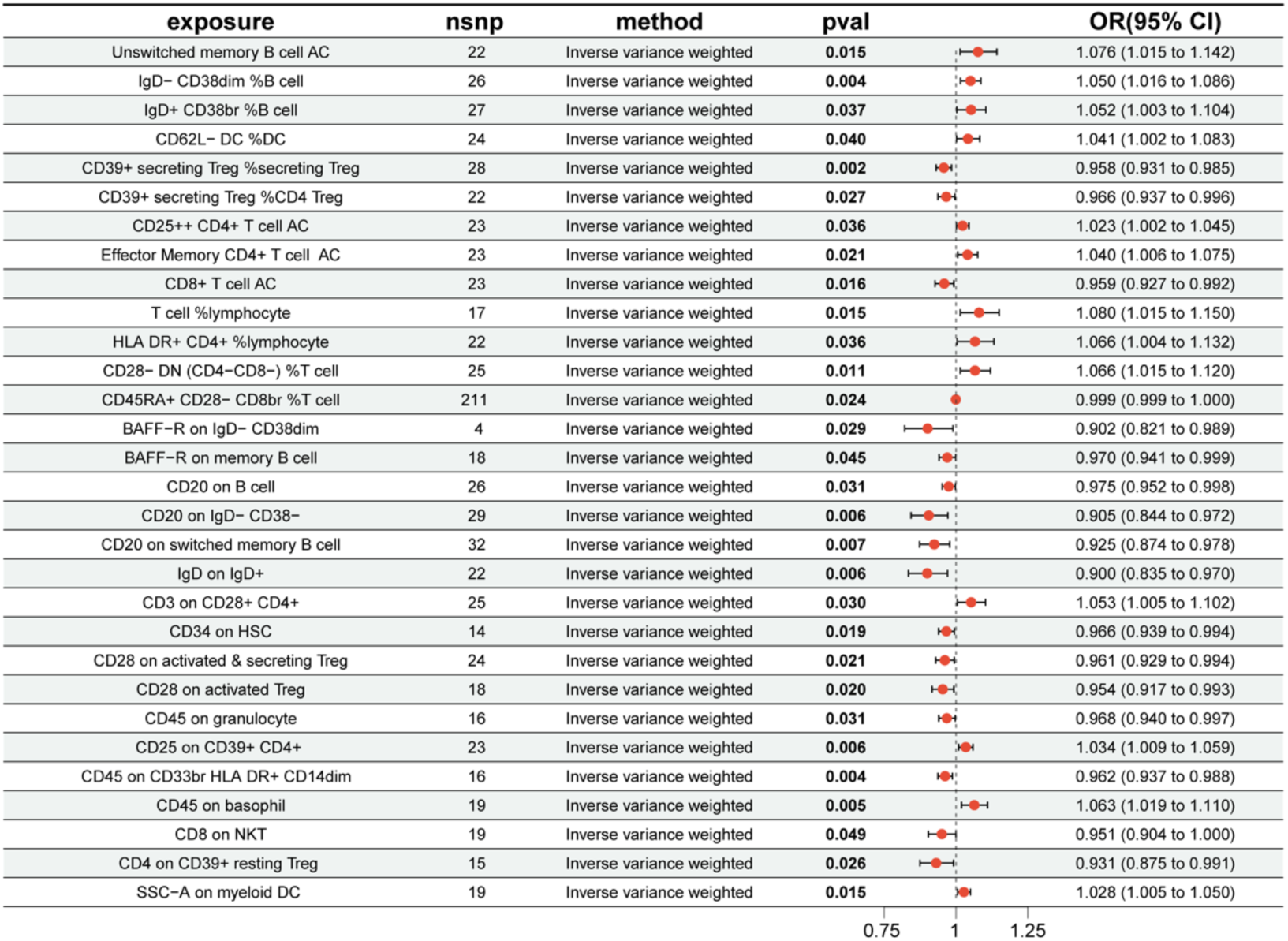
Forest plot for 30 immunophenotypes correlated with LC. MVMR study utilizing 731 immune cell phenotypes as exposures to investigate their potential causal effects on LC. 30 phenotypes were identified as significantly associated with LC. The OR and 95% CI for these associations are presented.

Leveraging these insights, we further examined the potential role of these immune cells as mediators in the pathway linking TB and LC. Significantly, our findings demonstrate that TB has specific causal effects on distinct immune cell subsets. This includes a reduction in CD4 on CD39^+^ resting regulatory T cell (OR= 0.916, 95%CI= 0.841–0.997, *P* < 0.05) (Figure 5). Moreover, there is no evidence supporting a reverse causal relationship between TB and immune cell (Table S8). The mediation analysis reveals that the pathway from TB to LC exhibit potentially mediated by CD4 on CD39^+^ resting regulatory T cell with 9.09% proportion (Table S9).

**Figure 5.**
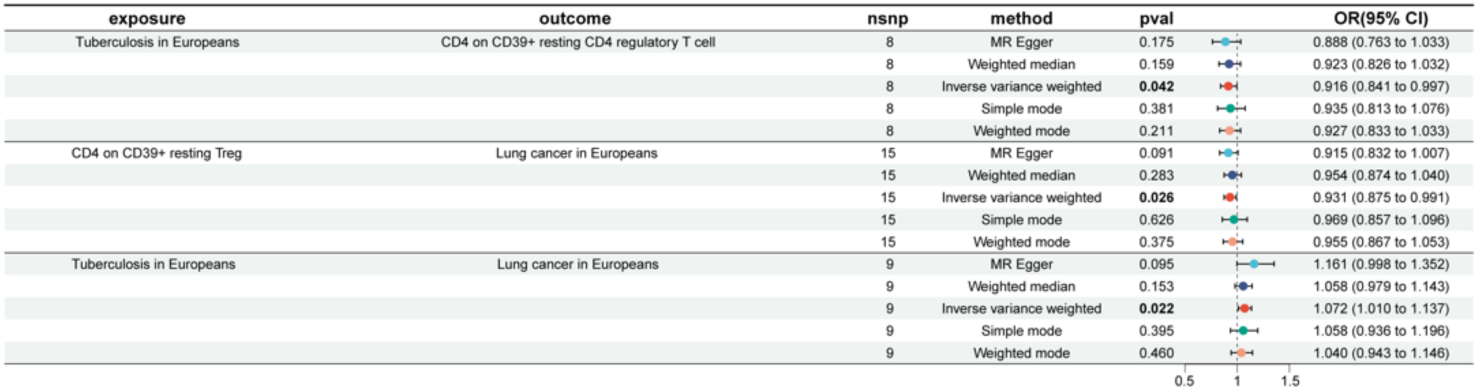
Forest plot: Mediation effect of TB on LC via immunophenotypes. MR analyses investigating the relationships between TB and CD4 on CD39^+^ resting CD4 Treg, and between CD4 on CD39^+^ resting Treg and LC. Analyses were performed using MR Egger, weighted median, IVW, simple mode, and weighted mode methods. The OR with 95% CI and *P*-values for each method are presented.

These results showed the consistent direction of the total, indirect, and direct effects, and that the LOO analysis supported the reliable causal relationship in the two-sample MR study of exposure to outcome, exposure to mediator, and mediator to outcome (Figure S6). This meticulous validation underscores the reliability of our results. This study not only underscores the complexity of immunological interactions but also enriches our understanding of the mechanisms at play in the co-morbidity of these diseases.

## Discussion

In the current study, we employed genetic predictors to investigate the causal relationship between TB and LC, while additionally exploring potential mediatory roles of immune cell phenotypes. Our findings demonstrate a unidirectional causal association from TB to LC Subsequent mediation analysis identified one immunophenotype—CD4 on CD39^+^ resting regulatory T cell—as a potential mediator in this pathway.

LC stands as a primary cause of cancer-related mortality globally, influenced by numerous contributing factors. Extensive epidemiologic literature explores the association between TB and LC. Notably, a retrospective cohort study involving 42,422 participants highlighted that LC risk was most pronounced within the first five years post-TB diagnosis, with a threefold increased risk persisting for over a decade[22]. Additionally, a meta-analysis of 32 studies indicated a significant elevation in LC risk following a TB infection[23]. Currently, several causal mechanistic studies elucidate this connection. TB patients often exhibit frequent DNA damage, cytokinesis defects, and an increased frequency of apoptosis and cell necrosis[24]. Additionally, the chronic inflammation caused by TB induces gene mutations in the lung parenchyma and repeated tissue damage, subsequently elevating the risk of cancer[25,26]. However, these observational studies are constrained by non-randomization, the absence of prospective and blinding methodologies, and the lack of large-scale validation in mechanistic investigations, thereby introducing potential confounding factors. To address these limitations, we applied MR to robustly assess the causal relationship between TB and LC. Our MR analysis utilized IVs comprising SNPs, which are randomly distributed and not influenced by external environmental factors[27]. Our bidirectional MR analysis in a European cohort identifies TB as a significant risk factor for LC development. Given the high prevalence of TB in underdeveloped regions of Asia, we further validated these findings using GWAS data from an Asian population.

To our knowledge, there is only one report demonstrating a causal relationship between TB and LUAD[28]. Our findings did not identify a significant association. This discrepancy may be attributable to several factors including differences in study design, population genetic diversity, or environmental co-factors. They used GWAS data from non-smoking Asian women, whereas we selected datasets from European and East Asian populations. Further studies are necessary to reconcile these findings and fully understand the genetic contributions to the various pathological subtypes of LC in various populations.

Extensive experimental evidence underscores the dual importance of not only mounting an effective immune response but also controlling it efficiently. Regulatory T cells (Tregs), which function by inhibiting the activity of other immune cells, play a pivotal role in preventing collateral tissue damage due to excessive inflammation in response to pathogenic bacteria[29]. Sakaguchi et al. elucidated that human CD4^+^ Treg cells encompass three phenotypically and functionally divergent subsets: CD45RA^+^Foxp3^lo^ resting Treg cells, CD45RA^−^Foxp3^hi^ activated Treg cells, and CD45RA^−^Foxp3^lo^ non-suppressive T cells, also referred to as non-Treg cells. The initial two subsets demonstrate suppressive capabilities *in vitro*, whereas the latter consists of cytokine-producing cells that lack suppressive functions[30]. In this study, utilizing a two-step MR analysis, we identified an immunophenotype—CD4 on CD39^+^ resting Treg cell—that may mediate the relationship between TB and LC. Intriguingly, subsequent analyses revealed a negative correlation between this immunophenotype and both TB and LC. CD39, also known as NTPDase1, is crucial for degrading extracellular ATP; its deficiency can trigger an excessive inflammatory response, leading to immune imbalance[31]. Such uncontrolled inflammation may provoke persistent tissue inflammation, ultimately fostering chronic inflammation that accelerates tissue destruction and predisposes to various tumors[32]. Genetically, our study predicts a causal link between TB and LC, identifying the potential intermediary role of CD4 on CD39^+^ resting Treg cell.

To our knowledge, this study represents the inaugural MR analysis investigating immune cells as mediators in the causal pathway between TB and LC. This work exhibits several strengths. Initially, GWAS data from European populations were employed to conduct causal analyses between TB and LC. Subsequently, these findings were corroborated and validated using population data from East Asian. Importantly, there is no potential overlap between the populations from these two regions, ensuring the independence of the samples. Moreover, we employed various complementary MR techniques to explore the causal links between TB, 731 immunophenotypes, and LC risk, thereby reducing the potential for residual confounding. Additionally, we conducted extensive sensitivity analyses to affirm the robustness of our conclusions.

This study has certain limitations. Firstly, our study focuses on the primary immune factors implicated in the pathogenesis of TB and LC, yet the detailed mechanisms of interaction remain to be elucidated. Our findings offer initial insights, which necessitate further validation through comprehensive *in vivo* and *in vitro* experiments at the cellular and molecular levels. Such investigations are essential to validate our initial findings and to potentially reveal therapeutic targets. Secondly, it is worthy to consider additional risk factors such as smoking habits, chronic lung diseases, and occupational exposures. These factors can significantly impact the development and progression of TB and LC, and their inclusion could provide a more comprehensive understanding of the etiology of diseases. Lastly, the MR results may be influenced by the heterogeneity of SNPs, introducing potential biases. Future research should aim to address these limitations by incorporating a more diverse array of genetic data and considering multi-factorial risk assessments to enhance the applicability of the findings.

## Conclusion

In conclusion, our extensive MR analysis determined that TB exerts a causal influence on LC, with no evidence of a reverse causal relationship. Based on these findings, we have identified an immune cell phenotype that may function as a mediator. This study provides novel evidence supporting a potential causal connection between TB and LC, highlighting immune factors as likely mediators. However, further corroboration of these results is necessary through additional extensive clinical investigations.

## Data Availability

All data produced in the present study are available upon reasonable request to the authors.

